# Wastewater-based reproduction numbers and projections of COVID-19 cases in multiple cities in Japan, 2022

**DOI:** 10.1101/2023.05.22.23290332

**Authors:** Shogo Miyazawa, TingSam Wong, Genta Ito, Ryo Iwamoto, Kozo Watanabe, Michiel van Boven, Jacco Wallinga, Fuminari Miura

## Abstract

**Background:** Wastewater surveillance has expanded globally to monitor the spread of infectious diseases. An inherent challenge is substantial noise and bias in wastewater data due to their sampling and quantification process, leading to the limited applicability of wastewater surveillance as a monitoring tool and the difficulty.

**Aim:** In this study, we present an analytical framework for capturing the growth trend of circulating infections from wastewater data and conducting scenario analyses to guide policy decisions.

**Methods:** We developed a mathematical model for translating the observed SARS-CoV-2 viral load in wastewater into effective reproduction numbers. We used an extended Kalman filter to infer underlying transmissions by smoothing out observational noise. We also illustrated the impact of different countermeasures such as expanded vaccinations and non-pharmaceutical interventions on the projected number of cases using three study areas in Japan as an example.

**Results:** Our analyses showed an adequate fit to the data, regardless of study area and virus quantification method, and the estimated reproduction numbers derived from wastewater data were consistent with notification-based reproduction numbers. Our projections showed that a 10-20% increase in vaccination coverage or a 10% reduction in contact rate may suffice to initiate a declining trend in study areas.

**Conclusion:** Our study demonstrates how wastewater data can be used to track reproduction numbers and perform scenario modelling to inform policy decisions. The proposed framework complements conventional clinical surveillance, especially when reliable and timely epidemiological data are not available.

## Introduction

The COVID-19 pandemic has presented a complex landscape for policymakers to navigate, with interfering factors such as vaccination, the emergence of new variants, and seasonality. Mathematical modelling has been employed by regional and national governments to monitor the disease in real-time, forecast epidemiological situations in near future (e.g., 1-2 weeks ahead), and inform policy decisions by projecting long-term trajectories under different scenarios [1,2]. Scenario modelling, exemplified by various research groups such as the COVID-19 scenario hub in the US and Europe [3,4], has contributed to more realistic and robust projections and a better understanding of epidemiological characteristics. However, accurate and standardized surveillance data are essential to capture temporal changes in disease dynamics, and it has become more challenging to obtain timely and unbiased epidemiological data via (passive) clinical surveillance due to changes in testing policies in many countries [5,6].

Wastewater surveillance has re-emerged as an alternative source of information during the COVID-19 pandemic [7,8]. Wastewater has the potential to monitor the prevalence by measuring virus concentrations excreted by infected individuals, which does not rely on patients’ symptoms or medical-seeking behaviour [8,9]. The effectiveness of wastewater monitoring has been demonstrated for various infectious diseases in the past [8,10,11], and the COVID-19 pandemic has accelerated its establishment in many countries [7,12]. Nevertheless, a remaining challenge inherent to wastewater surveillance is the substantial bias and noise in observed data due to the factors related to the sampling and quantification processes (e.g., higher water demand during daytime, dilution due to rainfall, PCR inhibition). To mitigate such biases, new molecular tools and sampling techniques have been developed [13]. Nevertheless, there remains the intrinsic noise in the observation process, and thus extracting true signals of epidemic growth requires data-analytic methods that can disentangle underlying trends from noisy data.

Previous studies have attempted to deal with the noise in wastewater data by using statistical or machine-learning based approaches [14–17]. The strength of these methods lies in the functional flexibility of models, which allows for the smoothing of noisy data (e.g., penalized splines [16], neural network [14,17]). These studies primarily focused on short-term forecasting and were successful in providing near real-time estimates [15,16]. However, a drawback of non-mechanistic models is that they do not necessarily provide biological interpretations, and thus the outputs from such analyses are difficult to use for policy guidance with further scenario analysis.

Mechanistic models have been applied to wastewater data in recent studies, with the primary aim of evaluating the predictive ability of models [18,19] or monitoring growth trends by computing effective reproduction numbers [20,21]. Yet, another important component, scenario modelling, has not been thoroughly explored in the combination of wastewater surveillance. Synthesis of multiple data streams would enhance the robustness of scenario modelling, and more importantly, there is a practical need to inform policymakers of strategic planning of interventions even in the absence of timely and reliable epidemiological data. In the current near-endemic situation of COVID-19, evaluating the potential impact of additional interventions such as vaccination campaigns is one of the key questions, even though notified data are not always fully available [5,6]. To this end, we need to exploit wastewater data and incorporate current transmission mechanisms (e.g., repeated infections related to emerging variants and waning immunity), which have not been explicitly captured in previous work [18,19].

In this study, we develop a modelling approach that accounts for reinfection and vaccination effects and propose a way to infer transmission parameters from wastewater data and integrate them with the scenario modelling framework. As a motivating example, we conducted wastewater monitoring in three municipalities in Japan between 2021-2022, where there was sufficient access to confirmation testing during the Omicron wave. We applied the proposed modelling approach to the collected data, first to evaluate its predictive performance and then to illustrate the impacts of different interventions (i.e., increasing vaccination coverages and reducing contact rates). Lastly, we discuss potential challenges that may arise when applying the proposed modelling framework to wastewater data for future implementations.

## Methods

### Wastewater data

We implemented wastewater surveillance in three study sites in Japan; Kyoto (population size: 778,000), Kanagawa (population size: 1,241,200), and City A (population size: 157,000). Wastewater samples were collected 2-3 times per week, and virus concentration in each sample was subsequently quantified with two different molecular methods, i.e. EPISENS-S and COPMAN [22,23]. The details of sampling methods and experimental procedures are provided in the **Supplementary Materials**. We normalized the observed SARS-CoV-2 concentration by a commonly used faecal indicator, i.e., Pepper mild mottle virus (PMMoV), to adjust for potential bias caused by sampling time and flow rate of influent wastewater. When the measured concentrations were below detection limits, we imputed them as 1 (copy/L) for computational convenience. We then constructed the time series of the normalized SARS-CoV-2 concentration by taking the geometric mean, and the data was used for further analysis.

### Epidemiological data

The number of daily confirmed cases within the same periods of wastewater was obtained from the corresponding local government websites (**Table S1**). As the coverage of the wastewater treatment plants does not always match the municipality areas, we calculated the daily number of cases in each catchment area by aggregating case data from multiple municipalities and weighting them by the proportion of the connected population size in each service area.

### Transmission model

We developed a compartmental SEIRS model to incorporate reinfections and viral shedding from infected individuals to wastewater, adapting the method of [18]. The disease states are: susceptible S(*t*), exposed but not yet infectious E(*t*), infectious I(*t*), and recovered R(*t*) (see the conceptual model diagram in **Figure S1**). The model considered reinfections among individuals who have been infected already, by defining the average duration of immunity 1/*ω* that was assumed to be 180 days (i.e., recovered transition back to the susceptible state at the rate of *ω*). We assumed fixed values for the mean latent period (1/*α*) and infectious period (1/*τ*) throughout the analysis. These parameters were defined based on the literature, and the details are provided in **Table S1** and **Table S2**. Two other parameters, the mean duration of virus shedding (1/*γ*) and the time-varying transmission rate (*β*(*t*)), were estimated by fitting the model to daily cases and/or wastewater data.

### Model fit

To estimate parameters, we converted the above (deterministic) model to a stochastic model and used an extended Kalman filter for the inference task. The Kalman filter or its extended filtering methods have been often used for calibrating a dynamic model with epidemiological surveillance data such as the daily number of reported cases [24,25]. In this study, we employed the filtering method to fit the transmission model to the observed daily cases, virus concentrations in wastewater, or both, by incorporating the observation errors in both data sources. The model dynamics, including the active virus shedding state A(*t*) and the (stochastic) observation errors, was described in the following system:

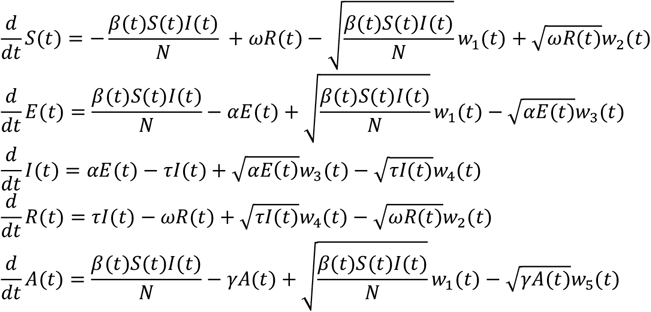

where the *w*_*i*_ are mutually independent white noise processes. We assume a closed population and thus *N* = *S*(*t*) + *E*(*t*) + *I*(*t*) + *R*(*t*). The transitions here are assumed to follow a binomial process, and the binomial distribution is approximated by the normal distribution (see details in [18]). The model outputs were then compared to the observed data (i.e., case data or wastewater, or both). Firstly, we assumed that the number of daily confirmed cases *y*_*C*_(*t*) is a fraction of infected individuals who newly become symptomatic on the date of observation

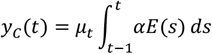

where *μ*_*t*_ is the reporting rate of newly confirmed cases out of the total number of infected cases on the observation day *t*. Since *μ*_*t*_ may change depending on the day of the week and the national holidays, the day-of-week effect were adjusted by estimating *μ*_*t*_ separately, and the holiday effect was further incorporated by reducing the reporting rate by 75% based on the observed maximum change in testing rates in Tokyo during December 2022 [26]. Secondly, the virus concentration in wastewater *y*_*w*_(*t*) is assumed to be proportional to the number of individuals shedding viruses *A*(*t*):

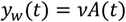

where *ν* is a scaling parameter specific to study regions.

### Effective reproduction number

To quantify the growth trend of an epidemic, the (instantaneous) effective reproduction number [27,28], the number of secondary infections caused by a single infected person at time *t*, is calculated. In this study, the effective reproduction number is obtained by the following equation:

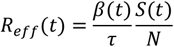

Where the transmission rate *β*(*t*) is obtained by fitting the model to either notified case data or wastewater data. To distinguish two different reproduction numbers, hereafter we use notification-based reproduction numbers 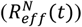 and wastewater-based reproduction numbers 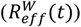 for further comparison. We computed the uncertainty in reproduction numbers by using estimates of *β*(*t*) and its standard deviation (SD) and visualized the uncertainty ranges of two standard deviations. As a reference to standard practice, we used the EpiEstim package [29] to estimate effective reproduction numbers from notified case data, assuming a serial interval is gamma-distributed with a mean of 3.5 days and a standard deviation of 2.4 days [30]. The EpiEstim estimators were then compared to the values computed by our approach.

### Forecasting and scenario projections

The model fitting via the Kalman filter allows an adaptive estimation of transmission rate *β*(*t*) at each time point, and thus we sequentially updated the estimated parameters using the most recent data points. To perform one-week ahead forecasting, we simulated daily reported cases over the next seven days using the most recent estimates of transmission rates and the number of individuals remaining in each state.

We examined two intervention scenarios; increasing vaccination coverage and reducing contact rates by non-pharmaceutical interventions (NPIs). Initial conditions for projections were determined using the estimated number of individuals in each state by fitting the model to the most recent observed data (**Table S1**). As a baseline scenario (i.e., a scenario without any additional intervention), we projected future cases in 4 months using the most recent estimate of the transmission rate *β*_0_.

In the scenario in which vaccination coverages are increased, the effect of additional vaccine uptakes was assumed to work as a transition from the susceptible to the recovered state (i.e., the vaccine mode of action was assumed to be “all-or-nothing” [31]). The transitioning proportion was calculated as *S*(*t*)(*c*_*vac*_ − *c*_0_)*VE*, where *S*(*t*) is the susceptible proportion, *VE* is the vaccine efficacy (assumed to be 60%), and *c*_0_ and *c*_*vac*_ are the vaccination coverages before and after the additional vaccination. The baseline vaccination coverage *c*_0_ was set as 70%, and we examined the expected impacts of increased coverage by varying *c*_*vac*_ as 80% and 90%. The effect of NPIs was modelled as a reduction in the contact rate, and thus the transmission rate after implementing NPIs was formulated as *β*_*NPIs*_ = (1 − ϕ)*β*_0_, where ϕ is the reduced ratio of contact rate compared to the baseline. In the main analysis the reduced ratio ϕ was set as 10%, and further reductions were examined in **Supplementary Text**. For both scenario analyses, we used the estimated baseline transmission rate *β*_0_ and its 2SD ranges as the uncertainty ranges of projections.

## Results

We implemented the wastewater surveillance system in three study sites in Japan. The observed wastewater data and the collected daily case data are illustrated in **Figure S2**. All data sources are provided in **Table S1**, and further details of two different RNA extraction/detection methods (i.e., COPMAN and EPISEN-S) are described in **Supplementary Text**. While there was a large degree of noise in individual observations of virus concentrations in wastewater, smoothed wastewater data indicated that growing and declining trends roughly matched with those observed in case data (**Figure S2**). This result was consistent regardless of the RNA extraction/detection method used or study areas.

The proposed modelling approach, using only wastewater data, described the epidemic trends in case data well at three study areas in Japan (**Figure 1**). Estimated parameters are listed in **Table S3**. In particular, the initial growth of epidemic waves in January 2022 was captured in a timely manner, which demonstrates the compatibility between notification-based and wastewater-based surveillance. Based on the estimated time-varying transmission rates and reporting rates, both total and reported cases were computed (shown in blue and red lines in **Figure 1**). The estimated ranges of reported cases were mostly consistent with the observed reported cases, and the large difference between the estimated total cases and the observed reported cases indicated that there may have been substantial under-reported cases around the peak of epidemic waves. By comparing different study areas, **Figure 1** illustrated that the uncertainty in estimates increased when the population size of the study area is small.

**Figure 1.**
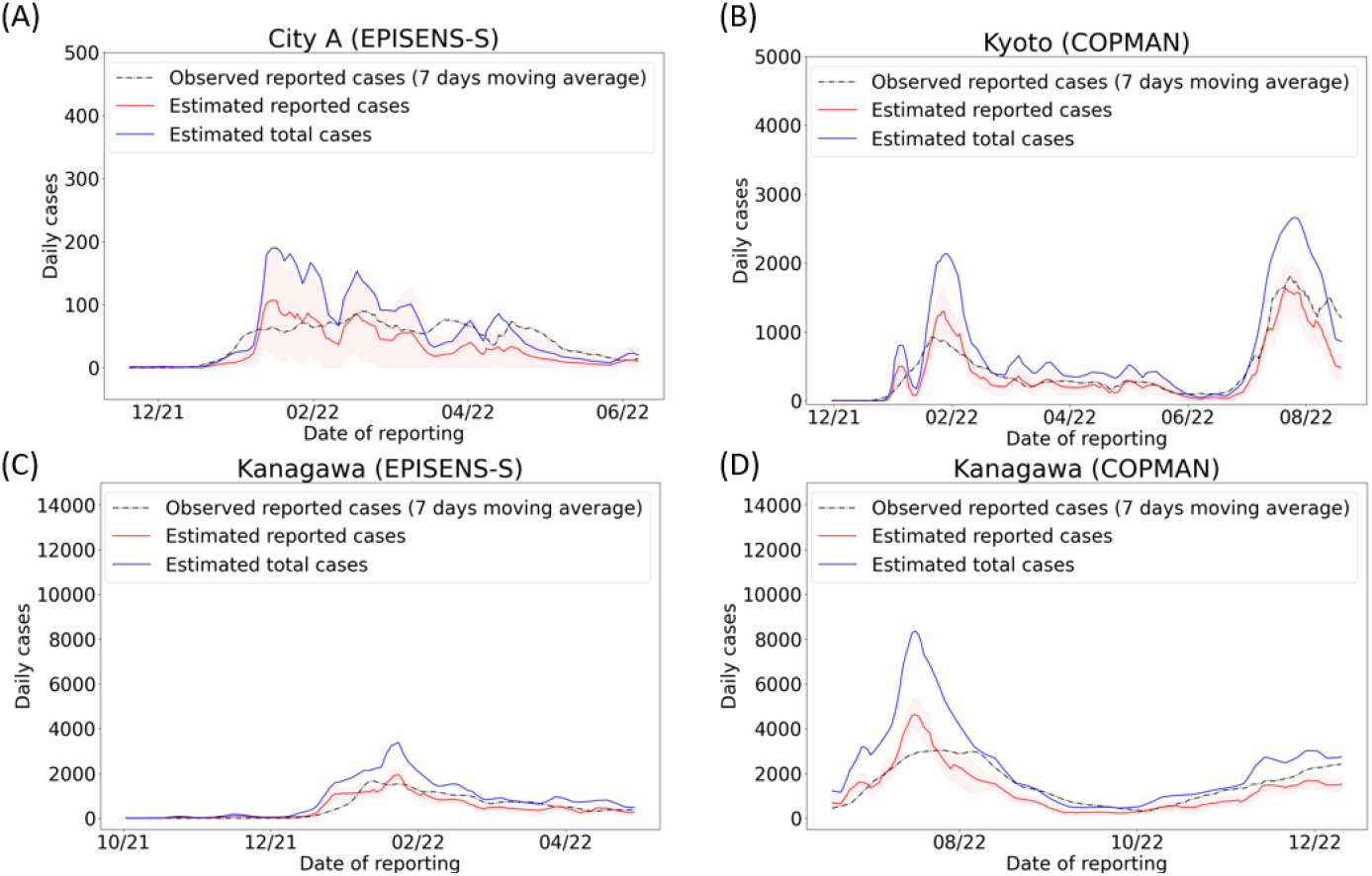
Estimated daily cases using only wastewater data in three areas in Japan. Black line indicates the observed daily reported cases, and red line and ribbon represent estimated daily reported cases with uncertainty bands of two standard deviations. Blue line corresponds to the estimated total cases, which was computed by incorporating under-reporting in the proposed modelling framework. COPMAN and EPISEN-S are different RNA extraction/detection methods, and the COPMAN method has a lower quantification limit of viral RNA.

To further validate our findings, we compared two effective reproduction numbers, i.e. notification-based reproduction number 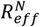 and wastewater-based reproduction number 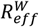 (**Figure 2**, shown in blue and red). This analysis showed that the computed 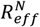 and 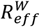 were comparable throughout the study period, suggesting that our modelling approach using wastewater data can provide a reliable proxy for tracking epidemic trends. Besides, this was further supported by the result that the computed 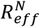 and 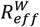 with our approach were mostly consistent with the values of a standard EpiEstim approach (**Figure 2**, shown in green). In general, however, the estimated values of 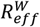 produced smoother curves with respect to time compared to the estimated 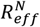. This indicated that the our approach with wastewater data alone may be less sensitive to abrupt changes in the epidemic, as the inherent noise in the data can hinder the identification of early signals.

**Figure 2.**
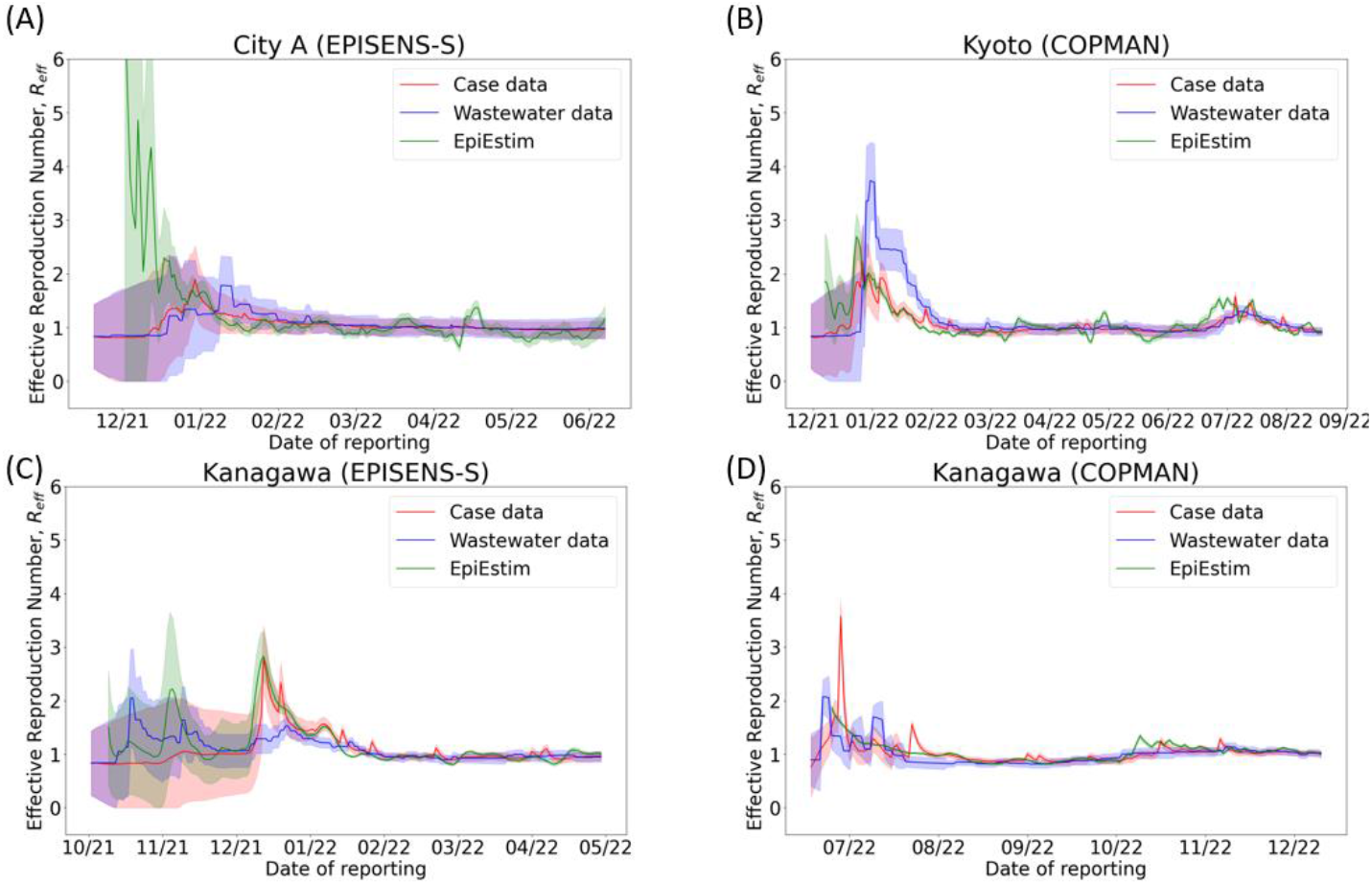
Estimated effective reproduction numbers using the proposed Kalman filter approach with only notified case data (red) and only wastewater data (blue), and using EpiEstim approach with only notified case data (green). Ribbons represent uncertainty ranges of 2 standard deviations (SD) for the proposed Kalman filter approach (red and blue) and 95% credible intervals for EpiEstim approach (green).

We conducted one-week ahead predictions of reported cases under three different conditions: using wastewater data only, case data only, and both wastewater and case data. To account for variations in observation frequency (two or three times per week), we aggregated daily case data over one week and compared the model predictions to the observed weekly number of cases. The prediction accuracy was evaluated by two error metrics (the Root-Mean-Square Error (RMSE) and the Mean-Absolute-Error (MAE)). **Figure 3** shows the one-week ahead prediction of weekly cases across different study sites and RNA extraction/detection methods, and the examined three conditions induced equivalent prediction abilities (**Table S4**). Interestingly, the model using both case and wastewater data did not necessarily show the best prediction performance, despite the utilization of all available data for the prediction. This result suggests that although wastewater data is suitable for capturing an epidemic trend, it may not necessarily improve the accuracy of short-term predictions.

**Figure 3.**
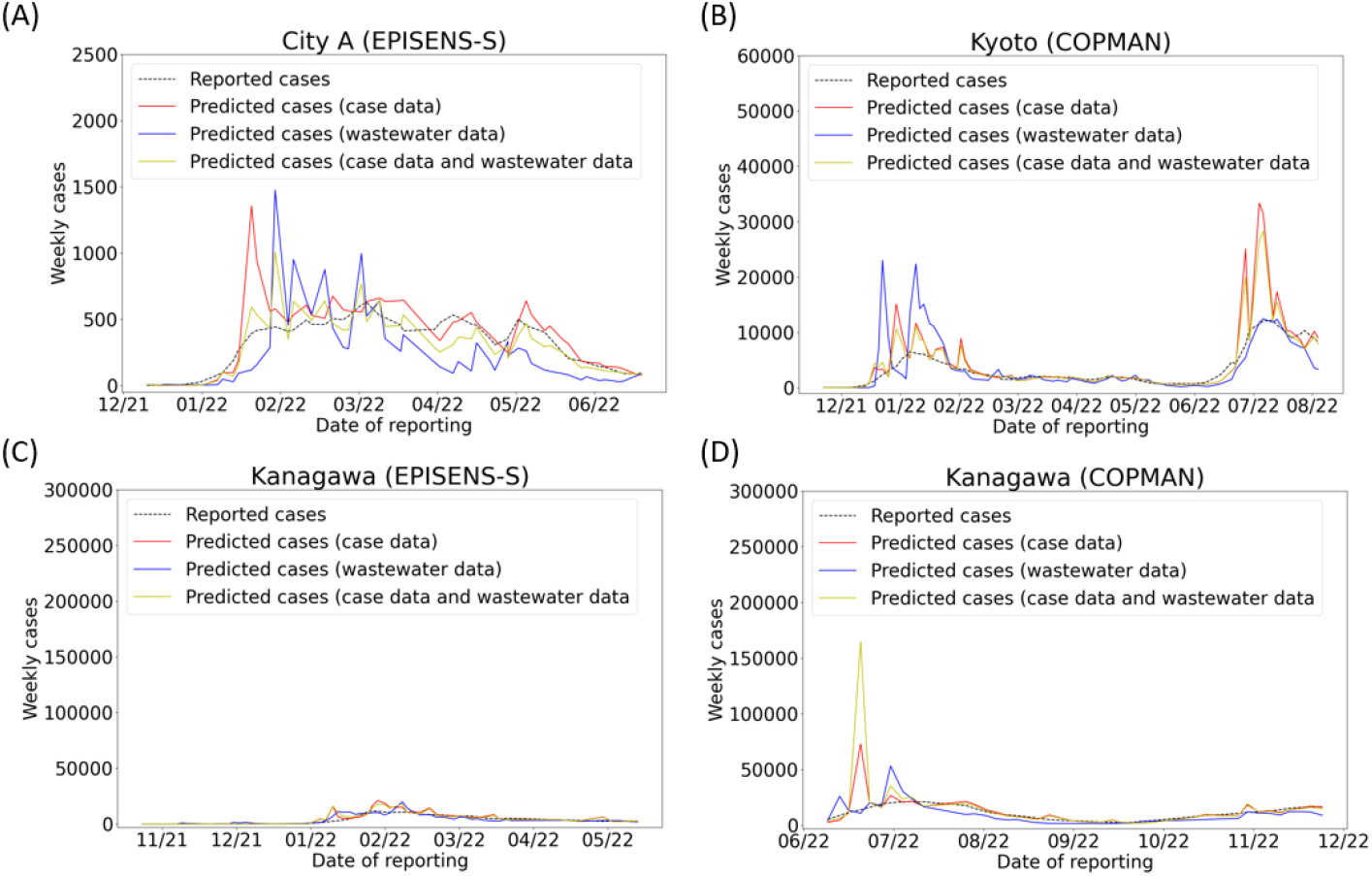
One-week ahead forecasting based on notified case data (red), wastewater data (blue), or both (yellow). The simulated number of cases over a week was adaptively updated via the Kalman filter and was compared to the observed reported cases (black).

To demonstrate model-based projections, we visualized the potential impacts of two different strategies (i.e., increased vaccination coverages and NPIs) in **Figure 4** and **5**. The forward simulations indicated that both strategies would expedite the decrease in daily cases when compared to the baseline scenario that imposes no additional interventions. While the projected baseline trajectories suggested an overall decreasing trend (green line in **Figure 4** and **5**), the uncertainty intervals in two study sites (Kanagawa and City A) indicated a possible increase in daily cases (green-shaded regions in **Figure 4(A)(B)** and **5(A)(B)**). The same trend in the baseline scenario can be more clearly seen in the projected cumulative cases (**Figure S3** and **S4**). We also performed more stringent NPI scenarios (**Figure S5-S8**); those scenario analyses showed an increase in the vaccination by 10-20% or a reduction in the contact rate by about 10% could alter the upper bound of the projected incidence into a declining trend in our simulation settings. Among the study sites, the largest reduction in projected cases was seen in Kanagawa during January-April 2023 where the incidence of cases was the highest (**Figure 5(D)**).

**Figure 4.**
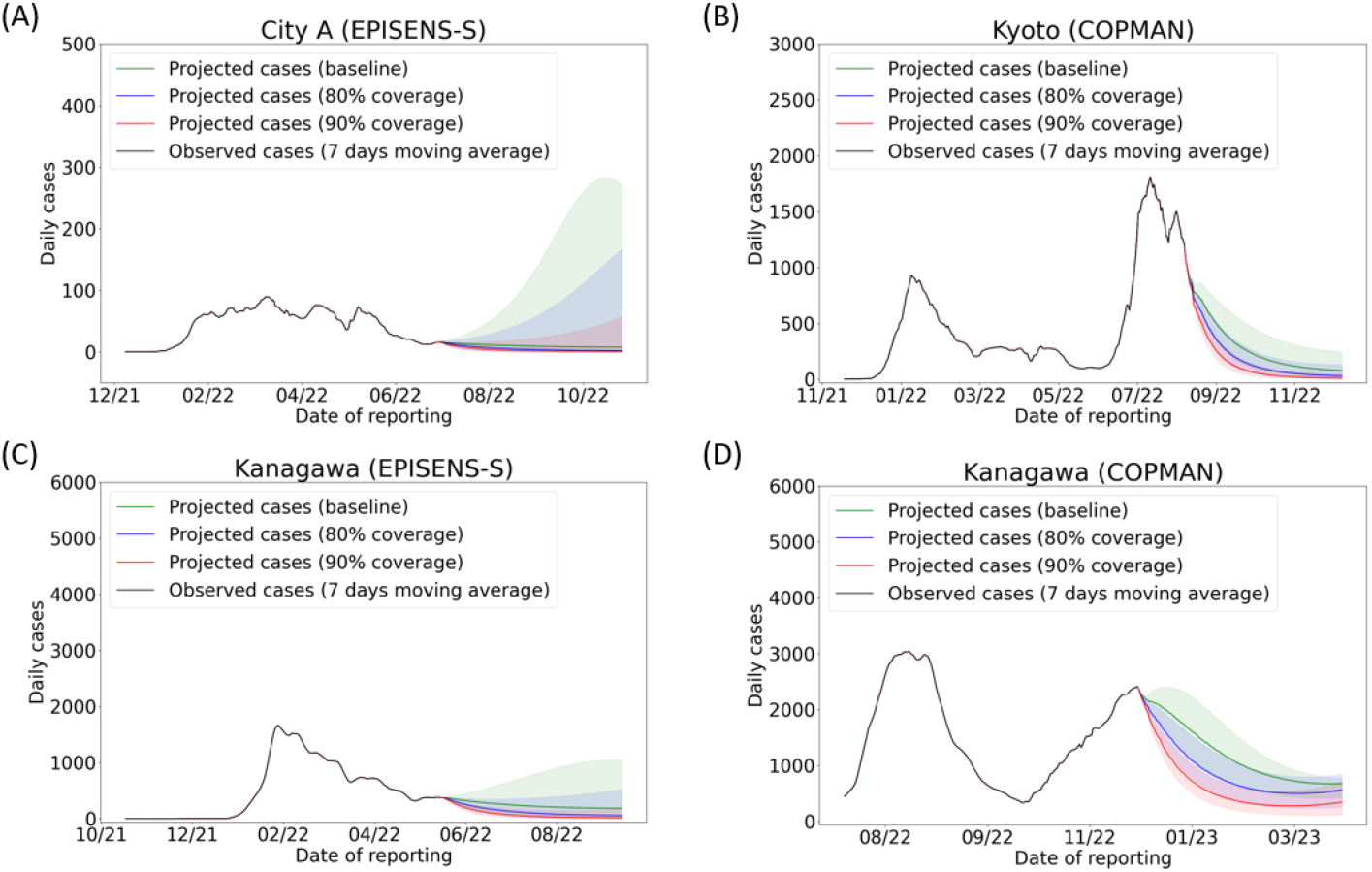
Model projected cases for increased vaccination scenarios. Vaccination coverages are set as 70% of the population for the baseline (green) and 80% (blue) and 90% (red) for scenarios with accelerated vaccine uptakes. Each ribbon represents uncertainty ranges of 2 standard deviations (SD) computed by the estimated variance of the baseline transmission rate.

**Figure 5.**
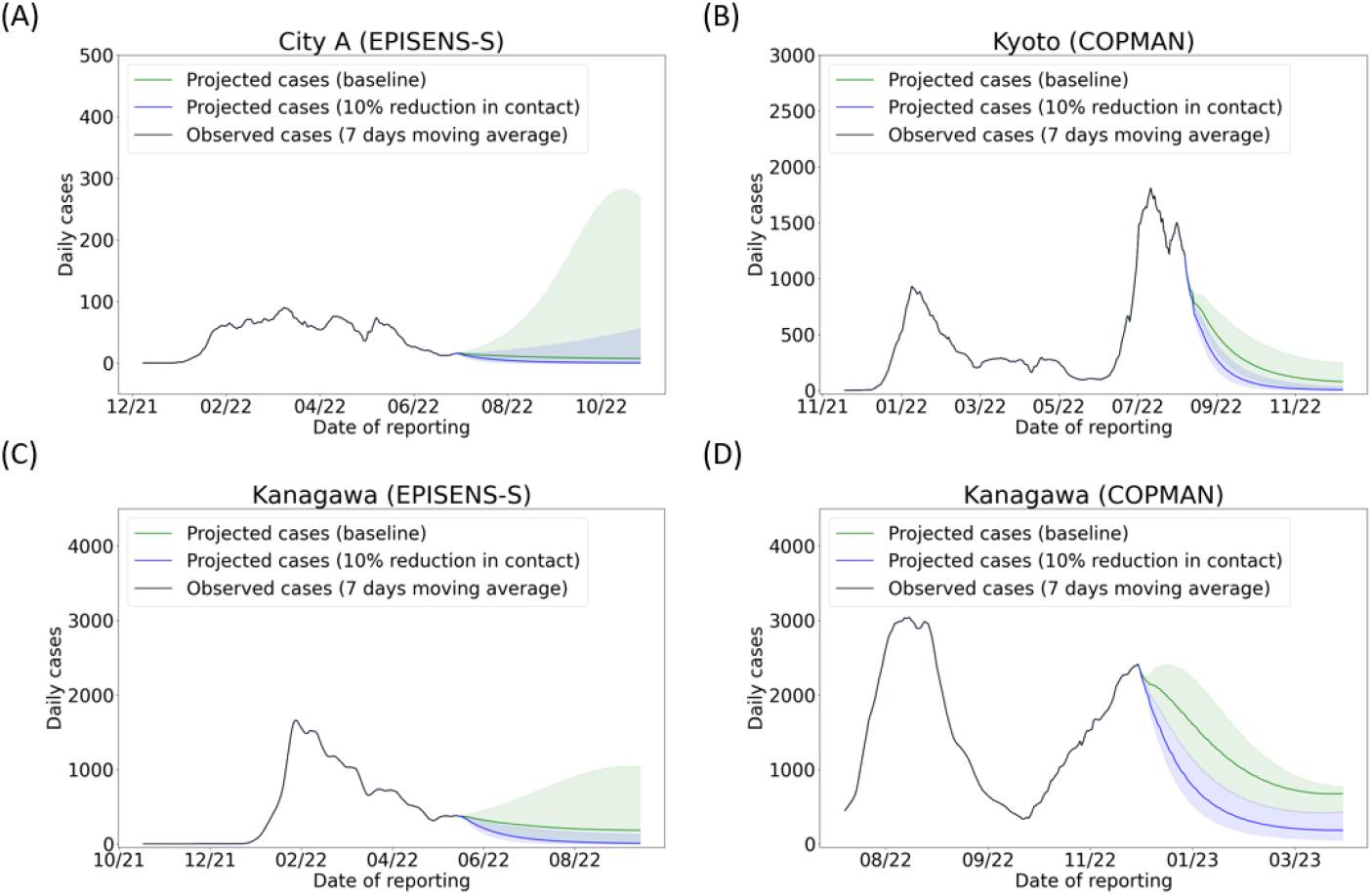
Model projected cases for a non-pharmaceutical intervention scenario. Relative contact rates are set as 1 for the baseline (green) and 0.9 (blue) and for scenarios with reduced contact rates. Each ribbon represents uncertainty ranges of 2 standard deviations (SD) computed by the estimated variance of the baseline transmission rate.

## Discussion

In this study, we showed that wastewater can capture the underlying trend of circulating SARS-CoV-2 infections and presented how scenario analyses can be provided to guide a policy decision by the proposed modelling framework. Our modelling approach translated the observed growth trend in wastewater data into effective reproduction numbers, which were consistent with estimated values by notified case data. As an application example, we further conducted scenario-based modelling analyses to illustrate the impact of different types of interventions on the projected number of cases. This highlighted the benefit of incorporating wastewater data into the current scenario modelling framework, regardless of the virus quantification method, especially when reliable epidemiological data are not obtainable.

The transmission model used in this study provided a good description of wastewater data (**Figure 1**). While previous literature in wastewater surveillance often claimed that machine-learning based models could capture more complex dynamics [14,17], our mechanistic model with parsimonious parameterizations yielded reasonable fitting results (**Figure 2** and **Table S4**). The main strength of our modelling approach is that all parameters have biological or epidemiological interpretations, and thus the outputs can be used for further scenario analysis. The interpretability and explainability are essential for informing policymaking as well as for (external) validity checks, in cases where there is a drastic change in transmission dynamics (e.g., the emergence of new variants).

Real-time monitoring of effective reproduction numbers via wastewater surveillance would be more effectively used if the notified case data suffer from substantial reporting delay or become less reliable. Effective reproduction numbers computed via case data are likely to capture the ‘true’ growth trend in an epidemic, regardless of the level of under-reporting, as long as the reporting rate is constant over the generation time (i.e., the average interval between the infections of an individual and its secondary case). Our analysis showed that wastewater-based reproduction numbers were consistent with notification-based reproduction numbers (**Figure 2**), suggesting that our approach can effectively monitor the epidemic trend via wastewater surveillance. Various methods have been proposed to compute effective reproduction numbers [29,32,33], and their limitations are widely discussed [27,34]. A common challenge is that those methods are prone to sudden changes in reporting system (e.g., case definition, testing policy, diagnostic capacity, etc.). By contrast, wastewater surveillance is more robust to such transition in the data collection process, and recently several approaches have been proposed to estimate effective reproduction numbers via wastewater data [18,20,21]. We proposed to extend the applicable range of this wastewater-based framework; reproduction numbers estimated by mechanistic modelling approaches, such as ours, would provide a coherent way to simulate possible trajectories of an epidemic by varying other parameters when the epidemiological situation is changing. This usability is important for the iterative policy-making process.

Using Japan as an example, we presented analyses by examining the impact of different intervention scenarios based on the proposed approach with the observed wastewater data. Our model projections showed that, in the case of two study cites (City A and Kanagawa) where the daily incidence was increasing, a 10-20% increase in the vaccination coverage or about 10% reduction in the contact rate may be sufficient to turn the epidemic into a declining trend. These scenario analyses are useful to understand how much additional effort would be needed for controlling the disease on average. However, if more granular scenarios and strategic planning are required (e.g., targeted interventions by age, occupation, etc.), additional epidemiological data would be essential, as wastewater data only captures an aggregated trend over the whole population in the catchment areas. Thus, wastewater surveillance should be used as a complementary tool to support the current epidemiological surveillance.

The present study provided insights for further improvement in wastewater surveillance and its applicability to scenario modelling. Our analysis suggested that the estimated growth trends via wastewater data were more consistent with case data when the prevalence was high and/or the population size covered by the sewage system was large (**Figure 1** and **2**). Conversely, when the prevalence is low, virus concentrations in wastewater would also become low and approach the detection limit, leading to uncertain RNA quantifications with larger variations. Although the sensitivity of molecular methods has been extensively discussed [8,35], the minimization of variations in observations (e.g., experimental errors, variations in water sampling process, etc.) is also the key to capture the underlying epidemic trends. While it is possible to incorporate unobserved variations with various modelling approaches, such as the one proposed in this study, the implementation of experimental and sampling systems with reduced errors (e.g., flow-proportional composite water sampling [13]) would enhance the accuracy of wastewater surveillance and expediate more reliable scenario analysis.

Our scenario analysis should be interpreted with caution. First, our formulation simplified the dynamics, and consequently various pathogen/host factors (e.g., age-dependent contact rates, infectivity and immune-escape effect by variant, seasonality, etc.) were aggregated into the estimated parameters. In particular, the projected impacts of vaccination strategy may vary in practice, due to differences in the timing of vaccinations or differing waning rates by age. Although our aim was to illustrate the proposed framework by using collected data in Japan with minimal parameterization, the model assumptions and possible extensions in the structure, such as age stratification, need to be considered when more data are available. For the best practice of scenario modelling, we should always accommodate alternative candidate models and should not rely on a single model, and scenario analysis needs to be adaptively updated.

In conclusion, we have illustrated how wastewater data can be translated into intuitive epidemiological quantities such as total cases and reproduction numbers, and how we can use wastewater data as an alternative source of information for scenario modelling to inform future policy. The proposed framework with wastewater surveillance complements and maximizes the benefit of clinical surveillance, especially when reliable and timely epidemiological data is not available.

## Data Availability

Wastewater and aggregated case data and all codes used for analysis are available on GitHub (https://github.com/AdvanSentinel/AS-SEIRS).

## Ethical statement

Ethical approval was not needed for this study. SARS-CoV-2 RNA concentration data in wastewater do not contain any privacy sensitive information.

## Acknowledgements

The authors acknowledge Prof. Byung-Kwang Yoo and Prof. Ung-il Chung at Kanagawa University of Human Services for their support in obtaining permission for wastewater sampling. The authors thank the staff members of the WWTPs at Kanagawa Sewerage Works Foundation and the crisis management office at Kanagawa prefecture for their support and permission to sample the wastewater.

## Authors’ contributions

Conceptualisation: SM, FM. Data curation: SM, TSW, GI, RI, FM. Formal analysis: SM, TSW, GI, FM. Methodology: SM, TSM, GI, RI, MvB, JW, FM. Software: SM, TSM, GI. Validation: SM, TSW, GI. Visualisation: SM, TSW, GI. Writing – original draft: SM, TSW, RI, FM. Writing – review & editing: SM, TSW, GI, RI, KW, MvB, JW, FM.

## Supplementary Materials

### Supplementary Text

#### Wastewater analysis

Influent wastewater samples were collected from wastewater treatment plants in three study areas in Japan (see **Methods** and **Table S1**). The wastewater samples were collected in sterile plastic bottles via grab sampling and immediately transported to the laboratory. The samples were processed with the concentration method described below on the day of sample collection.

The collected wastewater samples were analyzed by EPISENS-S [22] and COPMAN [23] to quantify the SARS-CoV-2 and PMMoV RNA concentration in wastewater. Briefly, for EPISENS-S, total RNA was extracted from suspended solid formed via low-speed centrifugation at 3,000 *g* for 10 mins in the 40 mL sewage samples, using the RNeasy PowerMicrobiome kit on a QIAcube system (Qiagen, Hilden, Germany) to obtain a final RNA extracted volume of 50 μL. One-step RT-preamplification was performed using 13.5 μL of the RNA extract and the CDC N1 forward and reverse (2019-nCoV_N1-F and 2019-nCoV_N1-R) and PMMoV reverse (PMMV_RP1) primers (Table S1, S2 in the Supplementary Material of [22]) with the iScript™ Explore One-Step RT and PreAmp Kit (Bio-Rad Laboratories, Hercules, CA, USA). Thermal cycling conditions for RT-preamplification were as follows; 25°C for 5 mins, 45°C for 60 mins, 95°C for 3 mins followed by 10 cycles of 95°C for 15 s and 55°C for 4 mins. qPCR was performed with QuantiTect Probe PCR Master Mix (Qiagen, Hilden, Germany) in a total reaction volume of 25 μL containing 2.5 μL of pre-amplified products, which was performed using the primers and probe for SARS-CoV-2 (CDC N1) or PMMoV with a final concentration of 400 nM and 300 nM each, respectively (Table S1 and S2 in the Supplementary Material of [22]). Thermal cycling conditions for qPCR were as follows; 50°C for 2 mins, 95°C for 10 mins followed by 45 cycles of 95°C for 3 s and 55°C for 32 s. qPCR reactions were completed on ABI 7500 Real-Time qPCR system (Applied Biosystems), and the threshold value of relative fluorescent intensity (ΔRn) was adjusted to be 0.2.

For COPMAN, viruses were coagulated with the addition of 1 μL of polyaluminum chloride (PAC) followed by vigorous shaking for 30 times, and subsequent gentle shaking at 80–120 rpm for 10 min at 4 °C. The samples were then centrifuged at 3000 ×g for 10 min, and the supernatant was discarded. The samples were centrifuged again at 3000 ×g for 3 min and the remaining liquids were removed by pipetting. The debris was then transferred to a 1.5-mL tube and lysed with 250-μL SDS-based lysis buffer and digested by 14.25-μL proteinase K solution at 56 °C for 10 min. Crude RNA of 200 μL was extracted from the samples with phenol/chloroform/isoamyl alcohol (25:24:1), which was then purified with carboxyl-modified magnetic beads, to obtain a final RNA extract volume of 50 μL. An aliquot (2 μg or 13 μL) of the magnetic bead-purified total RNA was subjected to cDNA synthesis using the Reliance Select cDNA synthesis kit (Bio-Rad Laboratories) under the following conditions: 50 °C for 60 min, 95 °C for 1 min in 20-μL reaction mix with 2 pmol each of reverse primers of SARS-CoV-2, RSV, and PMMoV. The resultant cDNAs of SARS-CoV-2 and RSV were pre-amplified for 10 cycles by the Biotaq HS (Bioline Reagents Ltd., London, UK) under the following conditions: 95 °C for 10 min, and 10 cycles of 95 °C for 15 s, 55 °C for 15 s, and 72 °C for 30 s, in 30-μL volume reaction mix containing 9 pmol each of forward and reverse primers. PMMoV cDNA was not preamplified because PMMoV RNA usually exists in wastewater with high amounts. Finally, viral RNA was quantified from 2.5 μL of the preamp product for SARS-CoV-2 and RSV, and 2.5 μL of cDNA for PMMoV by qPCR using the TaqMan Environmental Master Mix 2.0 (Thermo Fisher Scientific) under the following conditions: 95 °C for 10 min, and 45 cycles of 95 °C for 15 s and 60 °C for 30 s, in 20-μL singleplex reaction mix containing 10 pmol each of reverse and forward primers and 7.5 pmol of TaqMan probe.

#### Alternative vaccination scenario

In additional analysis where vaccination coverages are increased, the effect of vaccines was assumed to work as a proportional reduction in transmission rates (i.e., the vaccine mode of action was assumed to be “leaky” [31]), and the transmission rate after additional vaccination is:

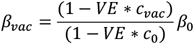

where *VE* is the vaccine efficacy (assumed to be 60%) and *c*_0_ and *c*_*vac*_ are the vaccination coverages before and after the additional vaccination, respectively. The baseline vaccination coverage *c*_0_ was set as 70%, and we examined the expected impacts of increased coverage by varying *c*_*vac*_ as 0.8 and 0.9. The results of projections are shown in **Figure-S3**.

**Figure S1.**
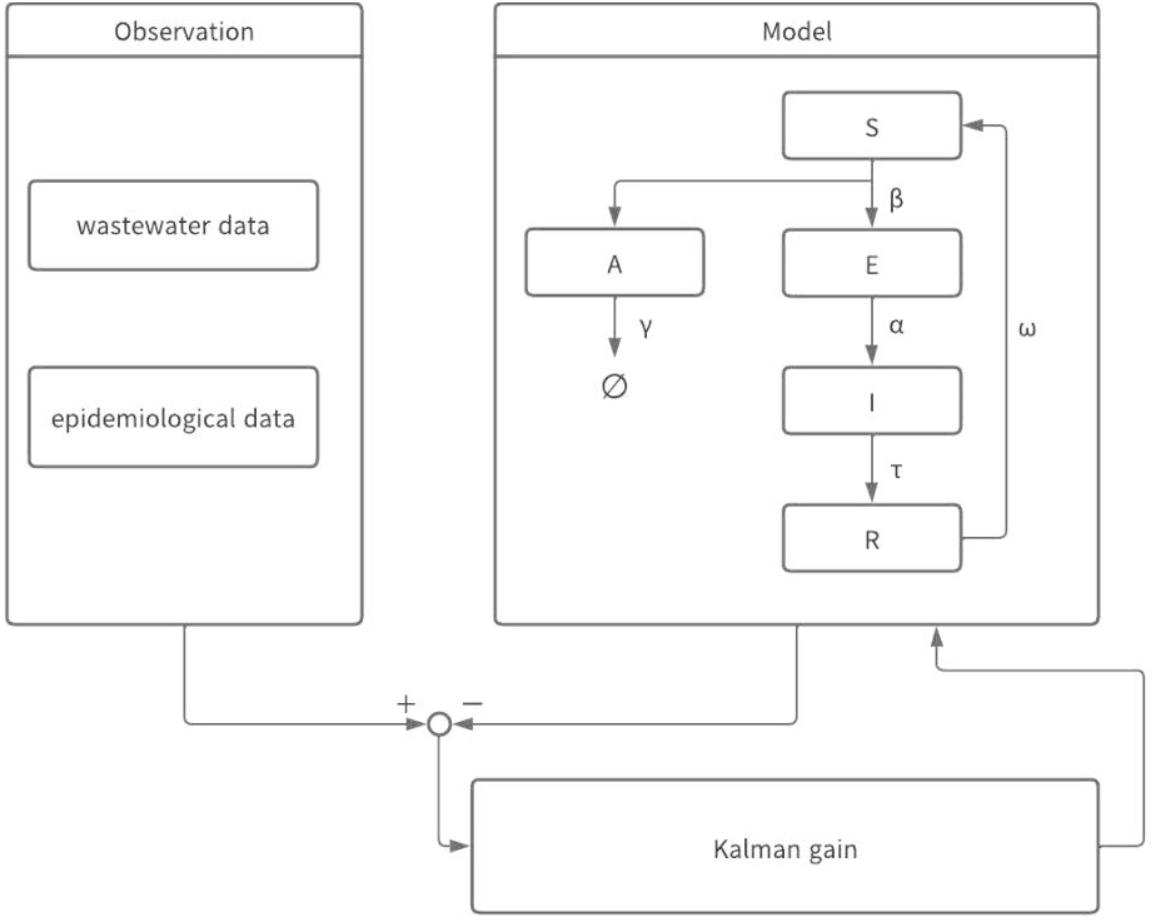
Schematic illustration of model structure.

**Figure S2.**
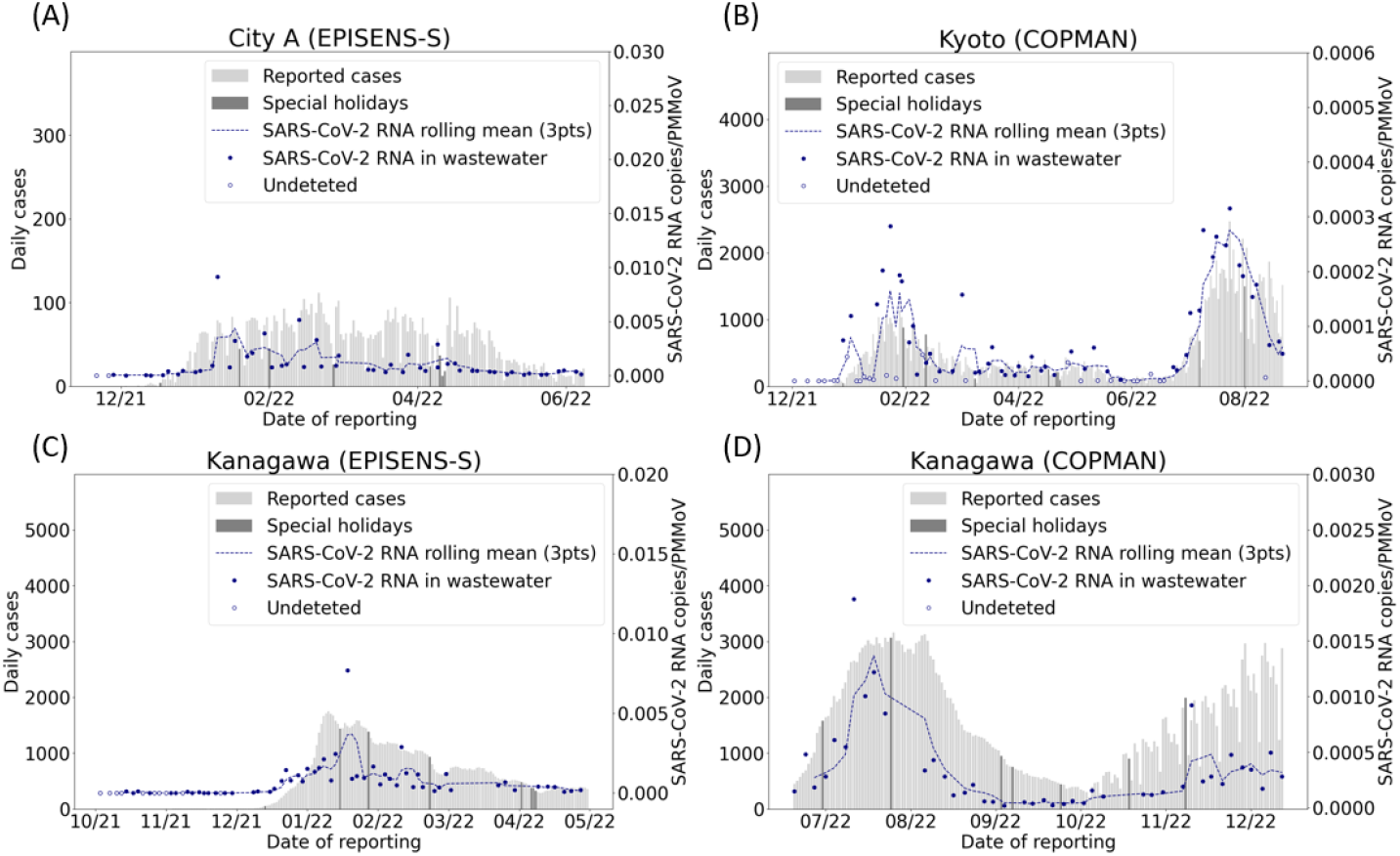
Collected wastewater and daily confirmed case data for different sites. Bars represent the daily number of reported cases, and darker bars indicate the cases reported on special holidays. Dot plots are SARS-CoV-2 RNA concentrations, and its rolling mean over three observed data points is shown as a blue line. White dots represent data points below the detection limit of RNA quantification methods.

**Figure S3.**
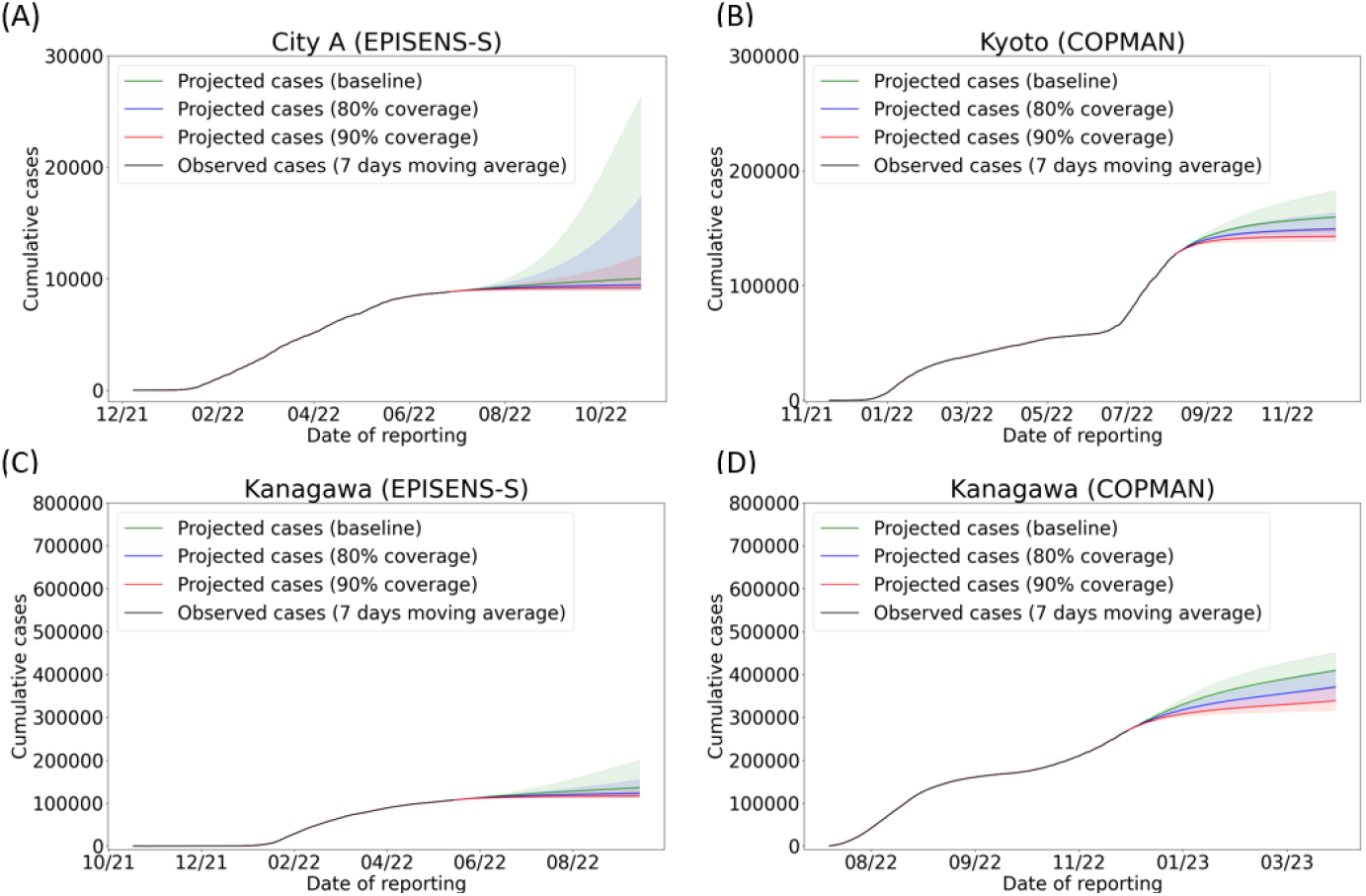
Projected cumulative cases in vaccination scenarios. Vaccine effect is assumed to reduce the baseline transmission rate proportionally (i.e., “leaky” effect). Vaccination coverages are set as 70% of the population for the baseline (green) and 80% (blue) and 90% (red) for scenarios with accelerated vaccine uptakes. Each ribbon represents uncertainty ranges of 2 standard deviations (SD) computed by the estimated variance of the baseline transmission rate.

**Figure S4.**
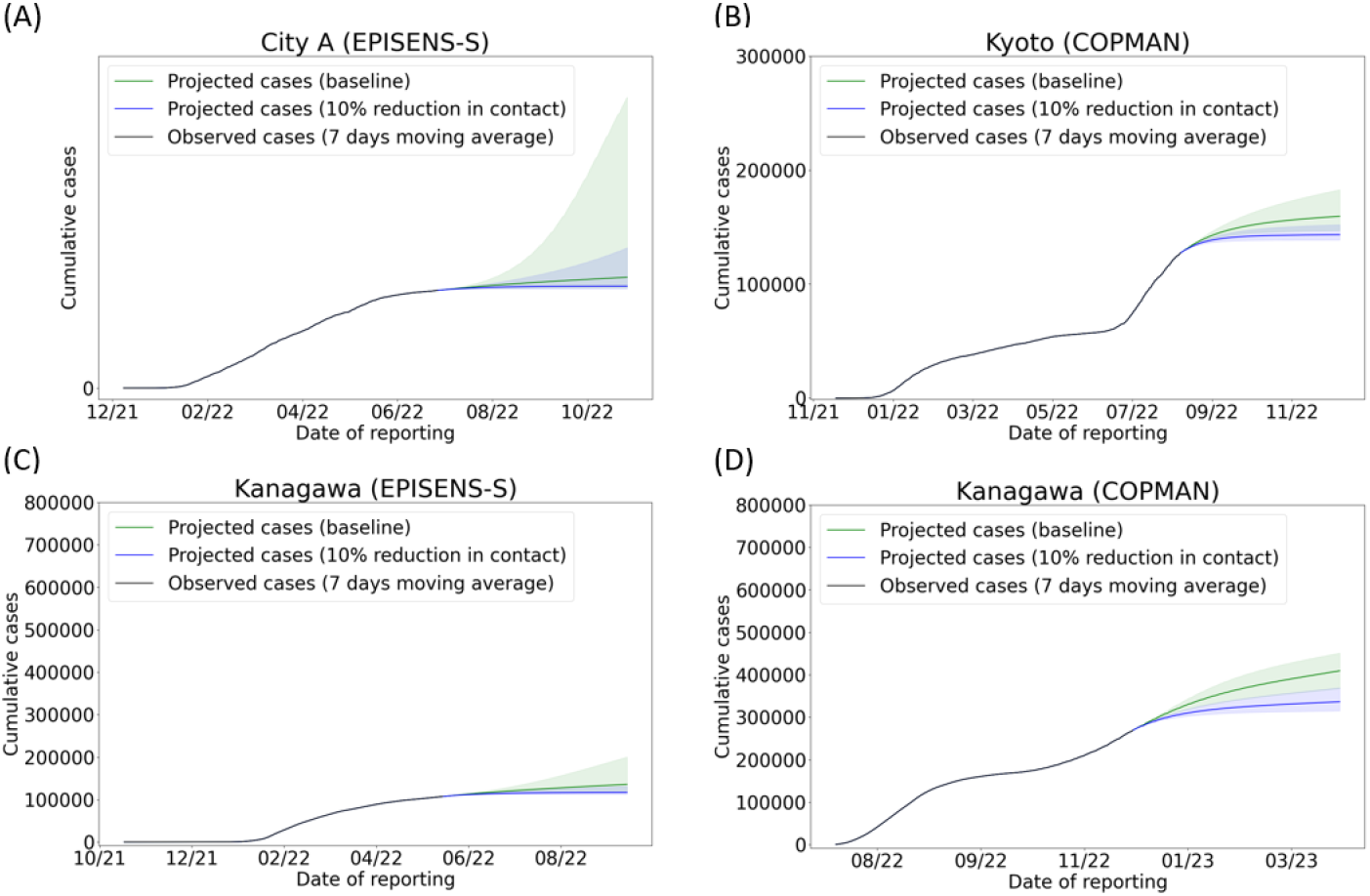
Projected cumulative cases in non-pharmaceutical intervention scenarios. Relative contact rates are set as 1 for the baseline (green) and 0.9 (blue) and for scenarios with reduced contact rates. Each ribbon represents uncertainty ranges of 2 standard deviations (SD) computed by the estimated variance of the baseline transmission rate.

**Figure S5.**
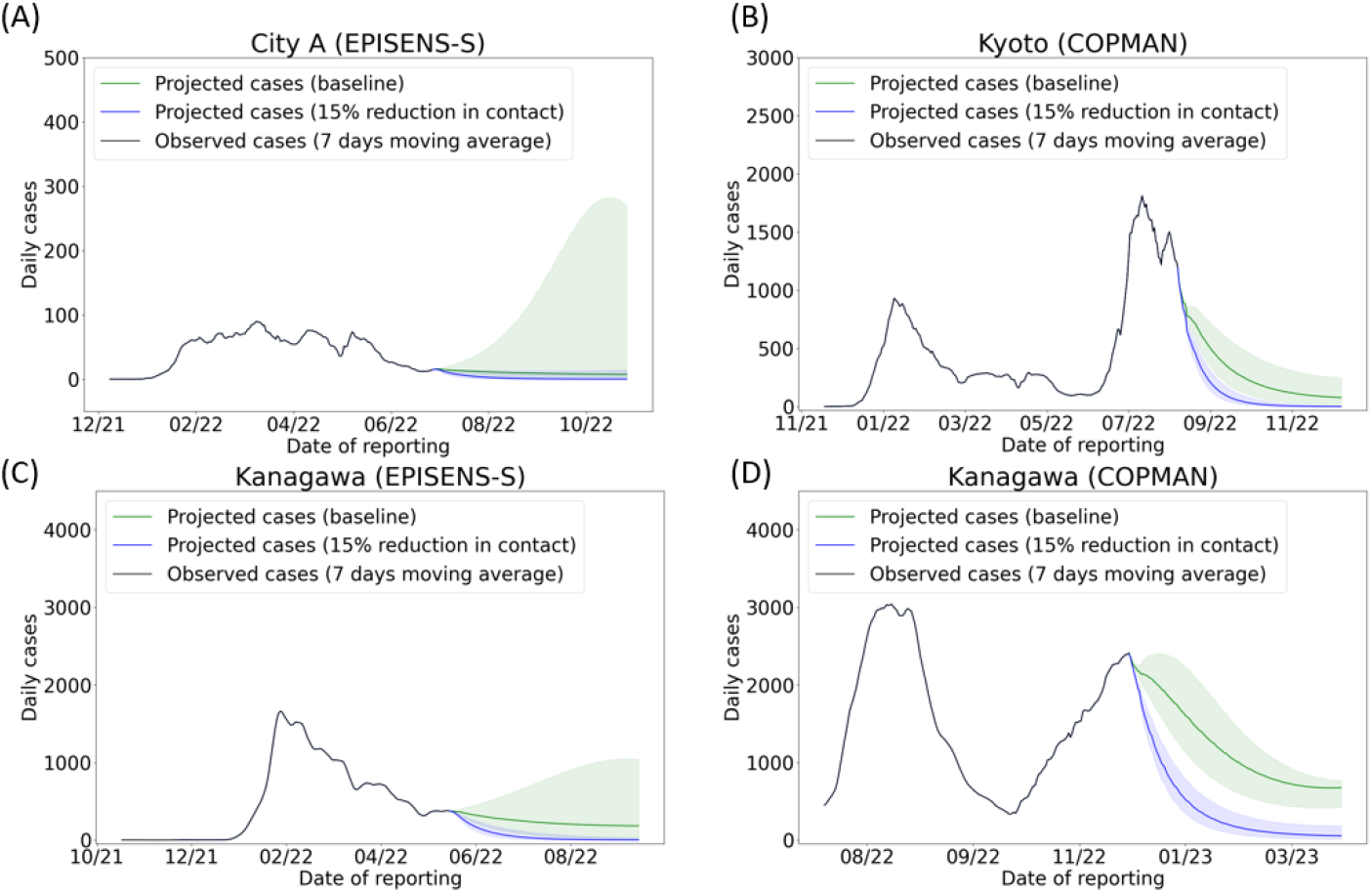
Additional analysis for a non-pharmaceutical intervention scenario. Relative contact rates are set as 1 for the baseline (green) and 0.85 (blue) and for scenarios with reduced contact rates. Each ribbon represents uncertainty ranges of 2 standard deviations (SD) computed by the estimated variance of the baseline transmission rate.

**Figure S6.**
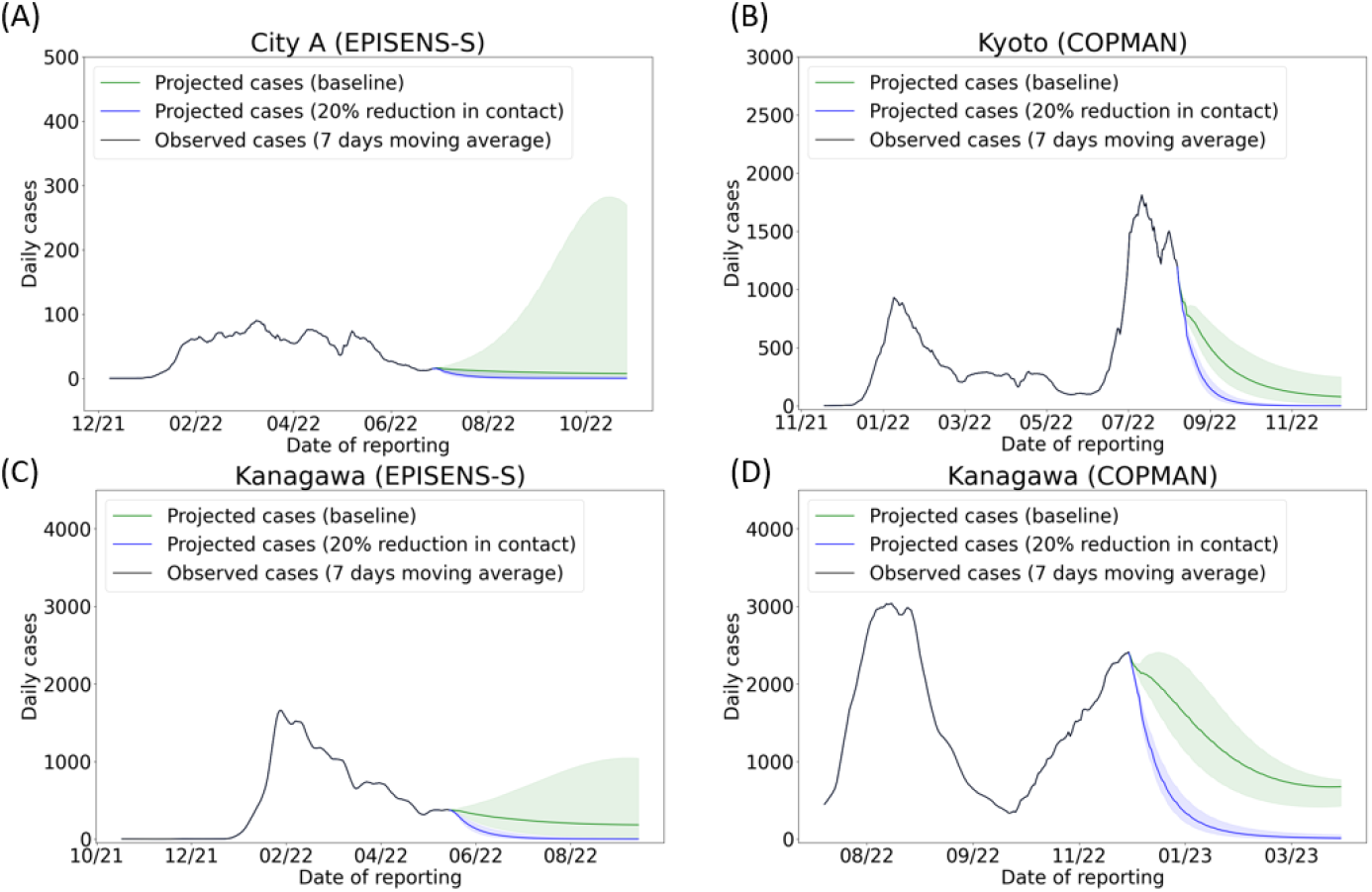
Model projected cases for a non-pharmaceutical intervention scenario. Relative contact rates are set as 1 for the baseline (green) and 0.8 (blue) and for scenarios with reduced contact rates. Each ribbon represents uncertainty ranges of 2 standard deviations (SD) computed by the estimated variance of the baseline transmission rate.

**Figure S7.**
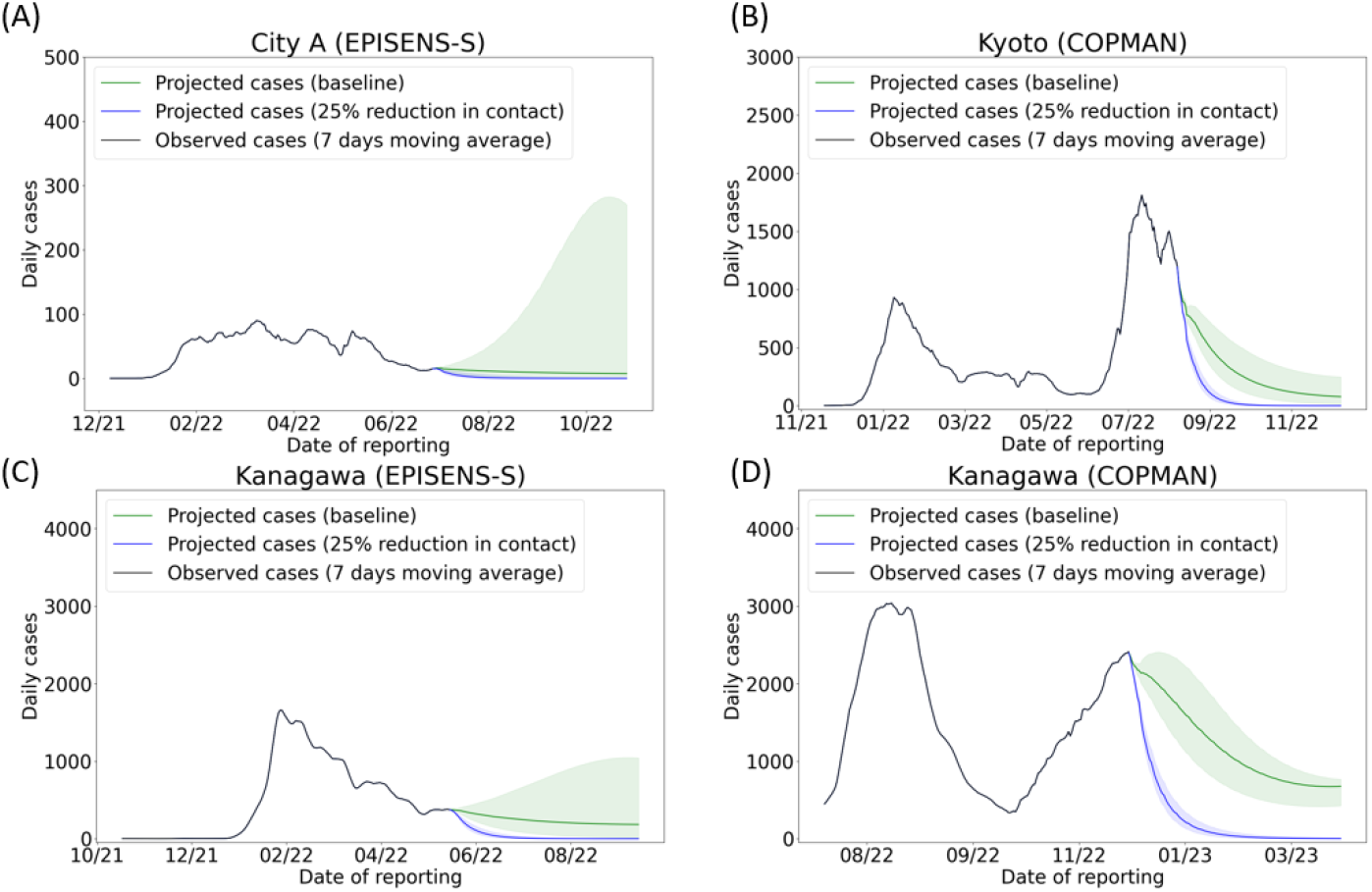
Model projected cases for a non-pharmaceutical intervention scenario. Relative contact rates are set as 1 for the baseline (green) and 0.75 (blue) and for scenarios with reduced contact rates. Each ribbon represents uncertainty ranges of 2 standard deviations (SD) computed by the estimated variance of the baseline transmission rate.

**Figure S8.**
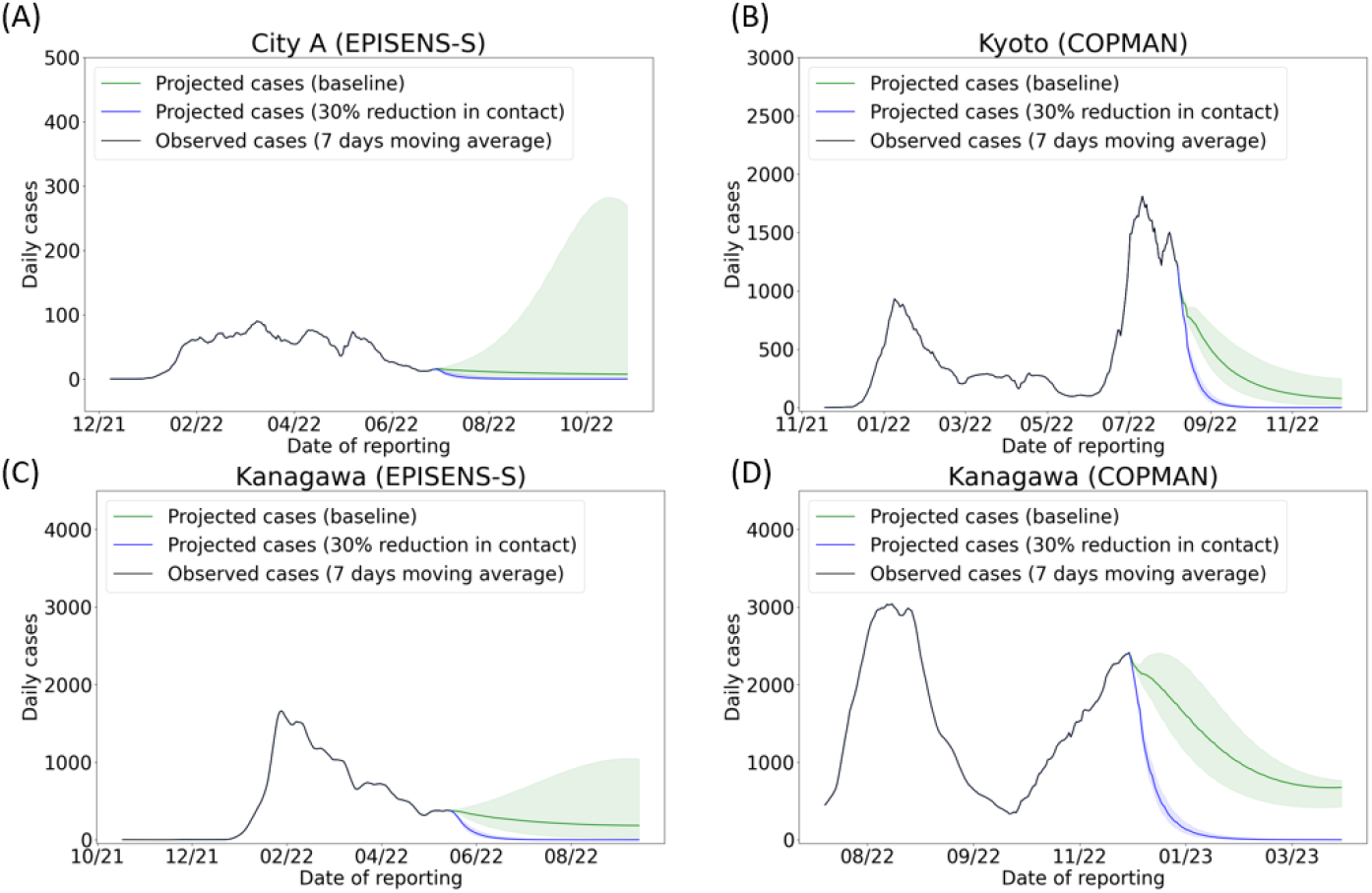
Model projected cases for a non-pharmaceutical intervention scenario. Relative contact rates are set as 1 for the baseline (green) and 0.7 (blue) and for scenarios with reduced contact rates. Each ribbon represents uncertainty ranges of 2 standard deviations (SD) computed by the estimated variance of the baseline transmission rate.

**Table S1.**
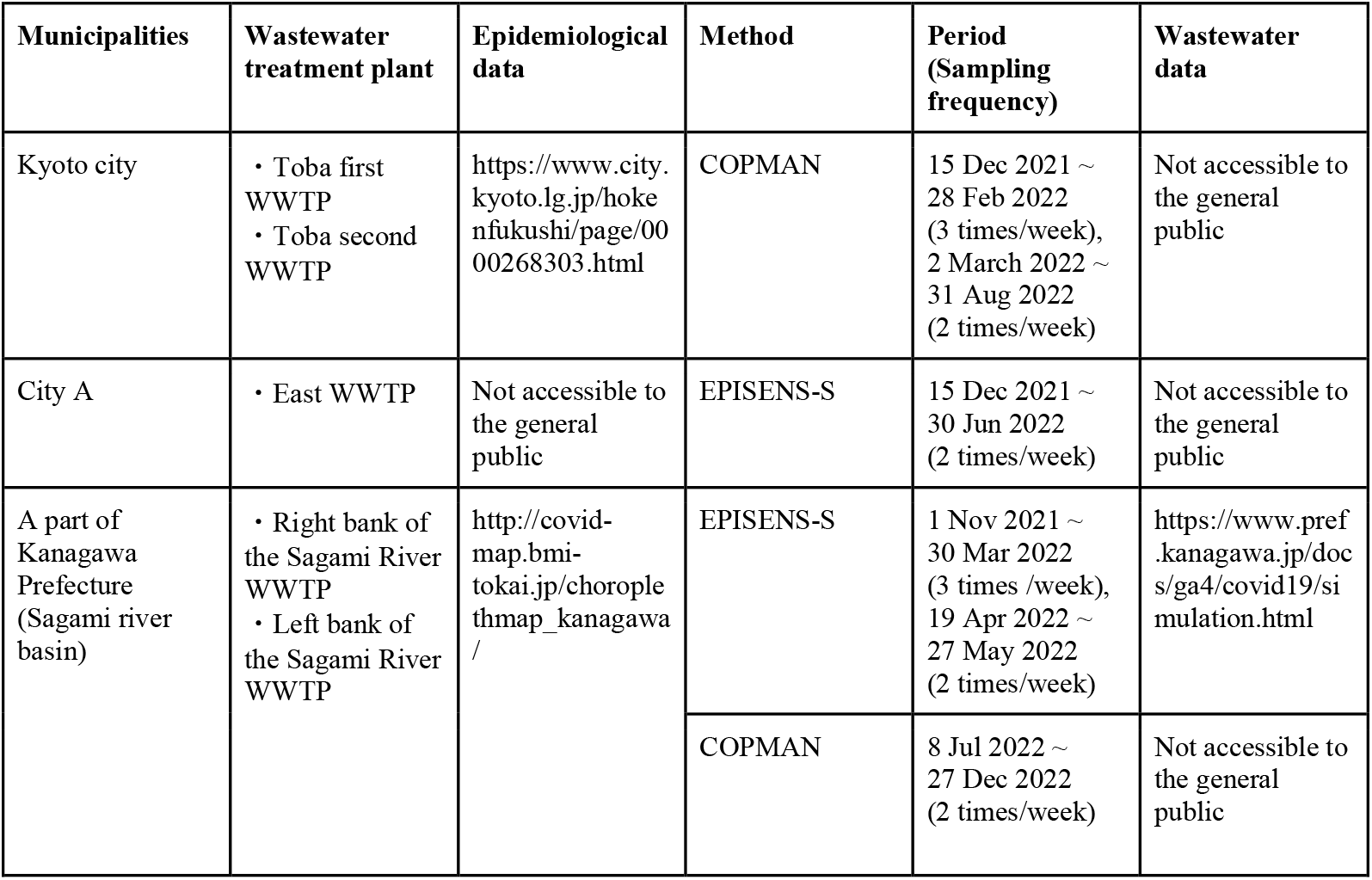
Data source.

**Table S2.**
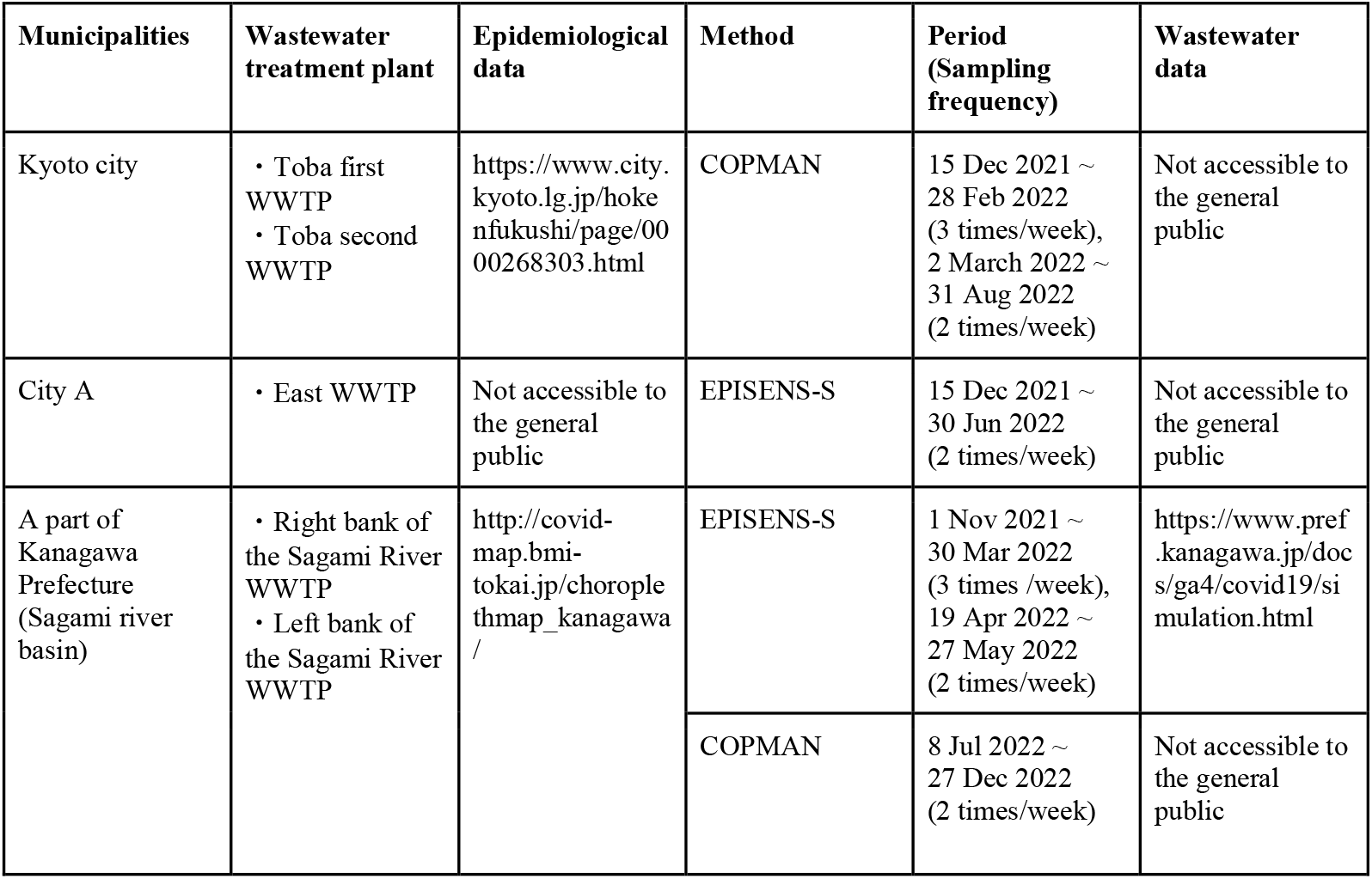
List of input parameters.

**Table S3.** List of estimated parameters.

| Parameter | Symbol | Kyoto city | City A | A part of Kanagawa Prefecture (EPISENS-S) | A part of Kanagawa Prefecture (COPMAN) |
| --- | --- | --- | --- | --- | --- |
| Shedding duration (days) | $1/\gamma$ | 1.97 | 0.64 | 1.71 | 1.74 |
| Scaling parameter | $\nu$ | $2.6 \times 10^{-11}$ | $2.0 \times 10^{-9}$ | $1.6 \times 10^{-11}$ | $1.7 \times 10^{-11}$ |
| Calibration period |  | 15 Dec 2021 to 31 Aug 2022 | 15 Dec 2021 to 30 Jun 2022 | 1 Nov 2021 to 27 May 2022 | 8 July 2022 to 27 Dec 2022 |

**Table S4.** Summary statistics for model fits.

| Municipalities |  | Kyoto city | City A | A part of Kanagawa Prefecture |  |
| --- | --- | --- | --- | --- | --- |
| Method |  | COPMAN | EPISENS-S | COPMAN | EPISENS-S |
| MAE* | Case data | 1869.3 | 91.6 | 3135.8 | 1303.3 |
|  | Wastewater data | 2032.9 | 178.1 | 4285.8 | 1397.5 |
|  | Case and wastewater data | 1475.9 | 81.8 | 5738.7 | 1305.9 |
| RMSE* | Case data | 4447.9 | 173.4 | 9632.6 | 2792.5 |
|  | Wastewater data | 3981.3 | 247.1 | 6969.1 | 2336.4 |
|  | Case and wastewater data | 3321.4 | 124.1 | 24071.8 | 2743.0 |

\*MAE and RMSE are calculated by comparing the predicted and actual numbers of cumulative new positive cases up to one week.

